# Strongly heterogeneous transmission of COVID–19 in mainland China: local and regional variation

**DOI:** 10.1101/2020.03.10.20033852

**Authors:** Yuke Wang, Peter Teunis

**Affiliations:** Center for Global Safe WASH, Hubert Department of Global Health, Rollins School of Public Health, Emory University, Atlanta, Georgia 30322, USA

## Abstract

**Background:** The outbreak of novel coronavirus disease 2019 (COVID-19) started in the city of Wuhan, China, with a period of rapid initial spread. Transmission on a regional and then national scale was promoted by intense travel during the holiday period of the Chinese New Year. We studied the variation in transmission of COVID-19, locally in Wuhan, as well as on a larger spatial scale, among different cities and even among provinces in mainland China.

**Methods:** In addition to reported numbers of new cases, we have been able to assemble detailed contact data for some of the initial clusters of COVID-19. This enabled estimation of the serial interval for clinical cases, as well as reproduction numbers for small and large regions.

**Findings:** We estimated the average serial interval was 4·8 days. For early transmission in Wuhan, any infectious case produced as many as four new cases, transmission outside Wuhan was less intense, with reproduction numbers below two. During the rapid growth phase of the outbreak the region of Wuhan city acted as a hot spot, generating new cases upon contact, while locally, in other provinces, transmission was low.

**Interpretation:** COVID-19 is capable of spreading very rapidly. The sizes of outbreak in provinces of mainland China mainly depended on the numbers of cases imported from Wuhan as the local reproduction numbers were low. The COVID-19 epidemic should be controllable with appropriate interventions.

**Funding:** No specific funding.

## Introduction

In December 2019, several cases of severe pneumonia appeared in Wuhan, the capital city of Hubei province in China. The outbreak was caused by a novel coronavirus: Severe Acute Respiratory Syndrome Coronavirus 2, or SARS-CoV-2 [1] and the disease (COVID-19) started to spread rapidly [2] in Wuhan. As of 19 February 2020, globally 75,204 cases have been confirmed with 2,009 deaths in 26 countries [3]. In China, 83·2% (62,031/74,576) of the confirmed cases were located in Hubei province and 60·4% (45,027/74,576) were located in the city of Wuhan, where the outbreak originated [4, 5]. The “pneumonia of unknown etiology” appeared in Wuhan from 8 December 2019. Many early cases have been reported to be linked to the Huanan (Southern China) Seafood Wholesale Market (hereafter referred to as “the Market”) [6]. By 2 January 2020, 41 initial cases were confirmed as COVID-19 [7].

As the novel coronavirus epidemic was spreading within Wuhan, the Chinese New Year/Spring Festival (25 January 2020), the most important holiday in China, was approaching. In 2019, 2.99 billion of people traveled by bus, train, and plane during 40 days around Chinese New Year [8]. Wuhan, with a population size of 11 million, is one of the four most important railway hubs in China. With billions of people travelling and lots of family and friends gathering, there was greatly increased risk of rapidly spreading this newly emerging infectious disease, nationally, and even globally. On 19 January 2020, the first confirmed COVID-19 case outside of Wuhan appeared in Shenzhen, Guangdong [9]. As of 23 January 2020, confirmed COVID-19 cases have been reported in 29 provinces of mainland China and nine countries and areas outside of mainland China [10, 11]. On the same day, the central government of the People’s Republic of China initiated a lockdown in Wuhan and two nearby prefectural cities, Huanggang and Ezhou, in Hubei province, to prevent spreading of the COVID-19 outbreak [12, 13]. However, as many cases had “escaped” from Wuhan before the lockdown, COVID-19 has spread to most provinces. The numbers of cases imported from Wuhan for different provinces heavily depend on their connectivities with Wuhan. At the same time, municipalities responded differently, regarding timeliness and adequacy of measures (including declarations of public health emergency, holiday extension, event cancelation, and surveillance using infrared thermometers in public spaces.

In this study, we used a method for analysing transmission patterns based on the serial interval between clinical cases of COVID-19 [14]. Based on dates of symptom onset and contact information for confirmed COVID-19 cases in Tianjin province, it was possible to estimate the serial interval distribution. With this information, it was possible to estimate numbers of new cases caused by any subject infectious with COVID-19, or their reproduction numbers during early stages of the outbreak in Wuhan. More importantly, we examined the heterogeneity in transmission among 30 provinces of mainland China and also among 20 cities in Guangdong province.

## Methods

### Data sources and assumptions

Various data with different levels of detail were collected from different sources.

**Table 1:**
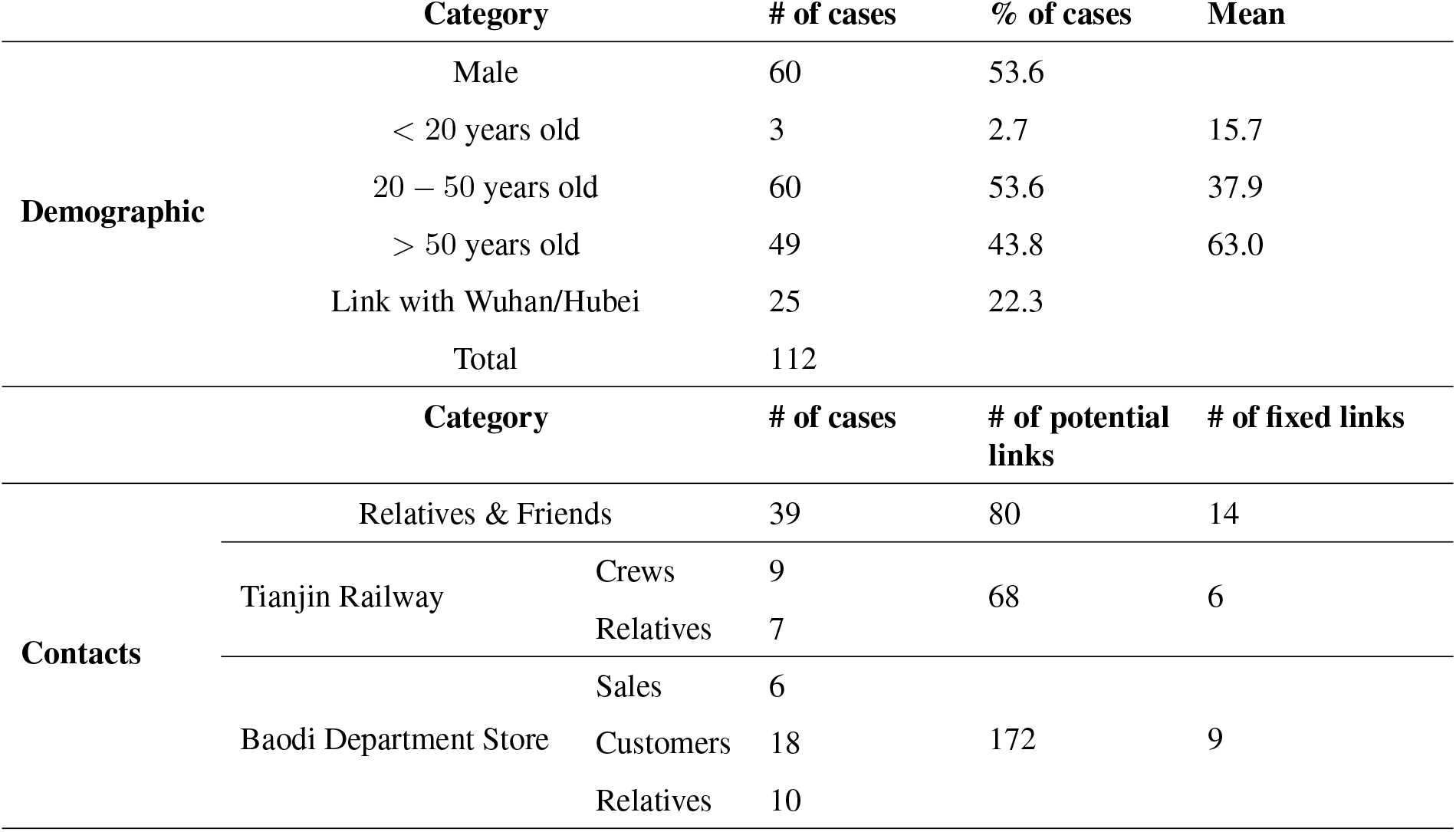
Characteristics of COVID-19 outbreak in Tianjin by 12 February 2020.

First, detailed data for 112 confirmed cases between 21 Jan 2020 and 12 Feb 2020 in Tianjin province was obtained from the website of Tianjin Health Commission, the website of Tianjin Centers for Disease Control and Prevention, and the official Weibo (China’s Twitter equivalent) of a local newspaper [15, 16, 17]. Collected information included demographic information, travel history to Wuhan/Hubei, date of symptom onset, and any contact information that could be found. See Table 1 for a summary.

Second, to evaluate the role of “the Market” in Wuhan and estimate the numbers of cases caused by contact with this source (its reproduction number) in early transmission, data for the first confirmed 425 COVID-19 cases with date of symptom onset and exposure information to “the Market” was extracted from a recently published report [6].

And in third place, we collected epidemic curve data: numbers of cases by province and date of confirmation from the Wikipedia page of COVID-19 cases in mainland China [18] and combined these data with detailed information, wherever available, about travel history and location (city and district) extracted from announcements of the Health Commissions of the provinces. These data were used to compare transmission of COVID-19 in different provinces and cities. Since the travel history to Wuhan is currently not available for all cases outside of Wuhan, the probability that any confirmed cases were linked with Wuhan was estimated using the 228 confirmed cases with known travel history between 20 January 2020 to 3 February 2020 in Beijing.

### Transmission analysis

Adopting terminology of Teunis et al.[14] a transmission probability matrix ***V*** can be defined where element *v*_*i,j*_ is the probability that subject *i* was infected by another subject *j*; ***v***_***i***_ is a vector of transmission probabilities linking case *i* to any other case. The total number of observed subjects is *n*. Elements of ***V*** can be estimated by utilizing a distance kernel *κ*_*i,j*_(*X*_*i,j*_ | *i* ← *j*), that defines a pairwise likelihood that subject *i* was infected by subject *j*. The distribution of the serial interval (Figure 1a): the distance in time between pairs of cases defines a practical distance kernel, translating the time intervals between symptom onsets in any two cases into a likelihood that these cases were linked as a transmission pair [19].

**Figure 1:**
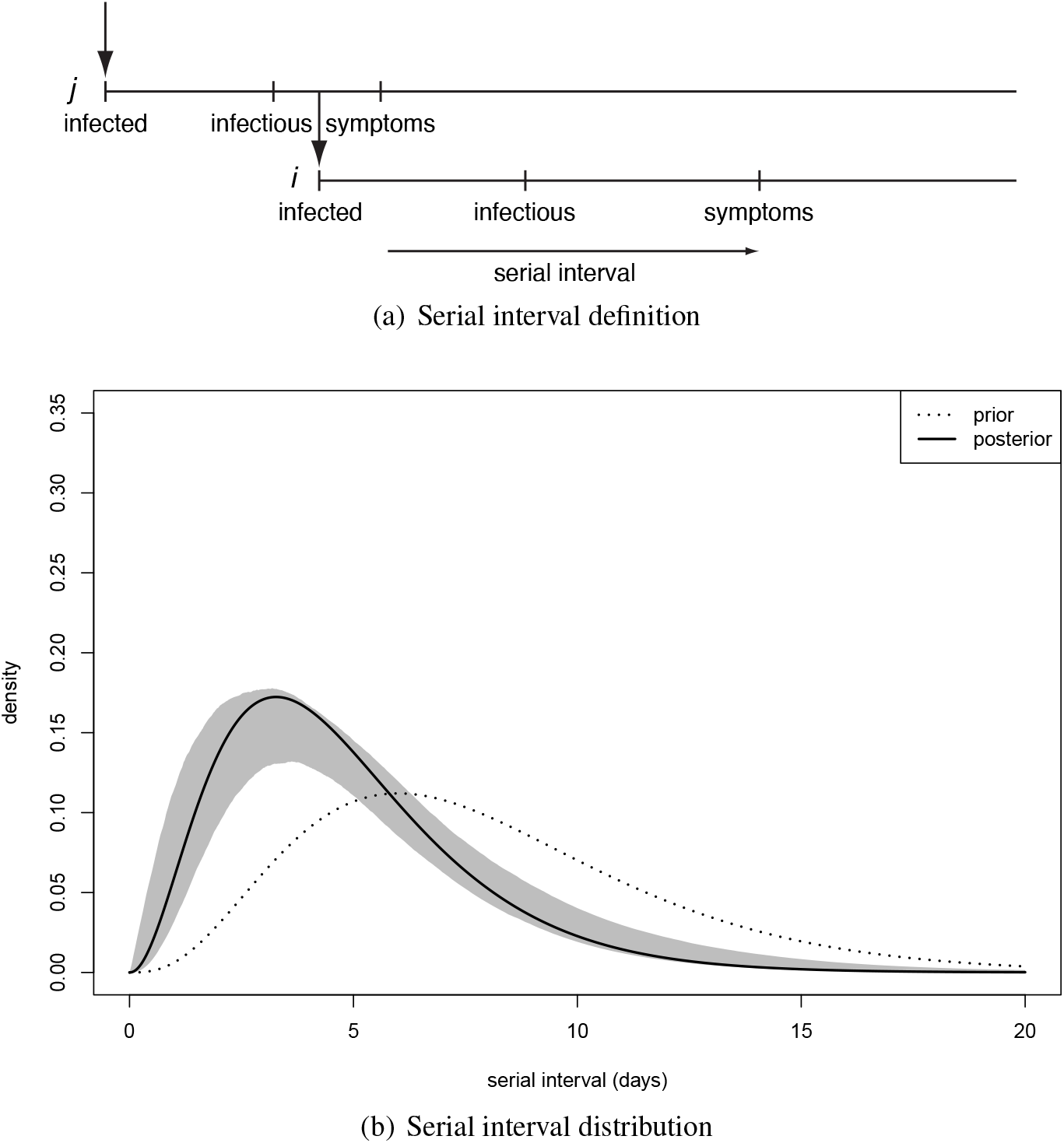
The serial interval between linked cases. (a) when a subject is infected, there is a latency until they start shedding virus and become infectious to others, then they (may) become symptomatic and can be detected clinically. When subject *j* infects subject *i* the time between symptom onset in case *j* and symptom onset in their descendant case *i* is the serial interval for onset of clinical symptoms. (b) Distribution of the serial interval for onset of clinical symptoms, estimated from COVID-19 clusters with partially known links (Figure2). Shown are the prior and the posterior mode, with 90% predictive interval.

As outlined in [14], the elements of the transmission probability matrix ***V*** may be estimated in a Markov chain Monte Carlo procedure. The elements of the transmission matrix are subject to constraints. Diagonal elements must be zero (subjects cannot infect themselves) and rows must add to 1 (unless the parent of the corresponding subject was not observed). Additional constraints may be imposed, for instance by preventing links between subjects known to have not been in contact. A mask matrix ***M*** (*n* × *n*, like ***V***) may be defined, with elements 1 where links are admissible, and 0 where they are not. This mask ***M*** may be applied to the matrix of kernels *κ*_*i,j*_() by elementwise multiplication. Elements of ***V*** representing pairs of subjects with inadmissible links are thus excluded: the corresponding *v*_*i,j*_ are set to zero, and they are not updated in MCMC estimation. The mask ***M*** can be used to define contacts: whenever subject *i* is known to have been in contact with another subject *j* element *m*_*i,j*_ = 1. And perhaps more importantly: when subject *i* is known to not have had contact with subject *k* then *m*_*i,k*_ = 0.

When a sufficient number of infectious contacts is known, the serial interval distribution may be estimated from outbreak data. First, the elements of the transmission probability matrix ***V*** are estimated, using (plausible) starting values for the parameters of the serial interval distribution. Then, ***V*** is fixed and the serial interval distribution parameters are estimated. Then the serial interval distribution is fixed and ***V*** is estimated. This alternating procedure can be repeated until no more improvement (in posterior probability) is found [14].

In the present analyses a special node (0) was defined, that has a uniform kernel *κ*_*i*,0_: any subject can be infected by node 0 at any time, within a given time range. Outside that time range *κ*_*i*,0_ = 0. Such a node could represent environmental transmission, i.e. from “the Market” to anyone in contact with that environment, or contact with a pool of infectious subjects, i.e. from any infectious subjects within Wuhan/Hubei to subjects outside of Wuhan/Hubei.

As the outbreak progressed, travel to Wuhan became increasingly less likely, especially after the lockdown of the city of Wuhan on 23 January 2020. However, contacts did not cease abruptly: travel records from the confirmed cases in Beijing showed a gradually decreasing probability of contact with Wuhan. Therefore the probability of linking any confirmed cases outside Hubei province to Wuhan was modelled as a logistic function of the date of confirmation

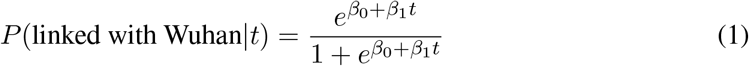

where *t* is the number of days after 20 February 2020. The parameters were estimated using the travel history records from the 228 confirmed cases in Beijing. The fitted logistic relation was then used to impute contacts with Wuhan sources for all cases outside Hubei province with unknown travel history.

Once infection has been imported from Wuhan, further transmission depends on local contacts: all cases cannot be assumed to be fully connected. Local contacts were imputed by distributing cases within a province or city into clusters. Cases within a cluster were assumed fully connected while cases in different clusters were not connected, thus allowing only transmission within a cluster and not between clusters. Cluster size was assumed random, with average size five people, and a high probability (88·8%) of small clusters (average 1 to 5 people) and a low probability (11·2%) of medium to large clusters (average 10 to 25 people).

The transmission probability matrix may be used to estimate reproduction numbers by calculating the row sums

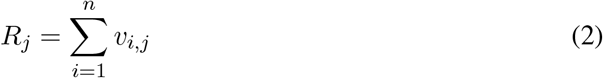

that represent expected numbers of cases infected by infectious case *j* (its outdegree), or its reproduction number. Reproduction numbers thus are calculated for individual subjects. Therefore they can be grouped in any way useful: by time, to illustrate the progression of transmission during the outbreak; by location, to illustrate spatial variation in transmission; or by other characteristics, like age or gender.

In order to translate the transmission probability matrix into a transmission network a multinomial sample of size 1 and probability vector row ***v***_*i*_ reduces the vector ***v***_*i*_ to a binary vector with exactly one element 1 and the rest zero. This reduced matrix may be interpreted as an adjacency matrix, equivalent to a directed graph representing a transmission tree of the outbreak [14].

## Results

For 112 confirmed cases in Tianjin between 21 January 2020 and 12 February 2020, the transmission network could be estimated based on the dates of symptom onset, augmented by contact information between cases. Table 1 shows some summary statistics of demographic and contact information for those cases in Tianjin. As contacts between many of the cases could be identified, so that many elements of the transmission probability matrix were known, joint estimation of the remaining unknown contact probabilities and the serial interval distribution was feasible. With a prior for the serial interval distribution set as Gamma(4,2), 1,000 updates were performed, alternating between probability transmission matrix and serial interval distribution parameters, with 100 iterations for each update and the best fit parameters were chosen as those with the highest posterior probability. Figure 2 shows an estimated (posterior mode) transmission network. Using the available contact information, there appeared to be considerable variation in the sizes of clusters of cases. There were two major clusters: one among crew members of Tianjin railway and another one among sales representatives and customers in Baodi department store. Figure 1b shows the posterior mode serial interval distribution as a gamma distribution (shape parameter 3·16, scale parameter 1·52), with a mean of 4·8 days. This best fit serial interval distribution has been used in all following analyses.

**Figure 2:**
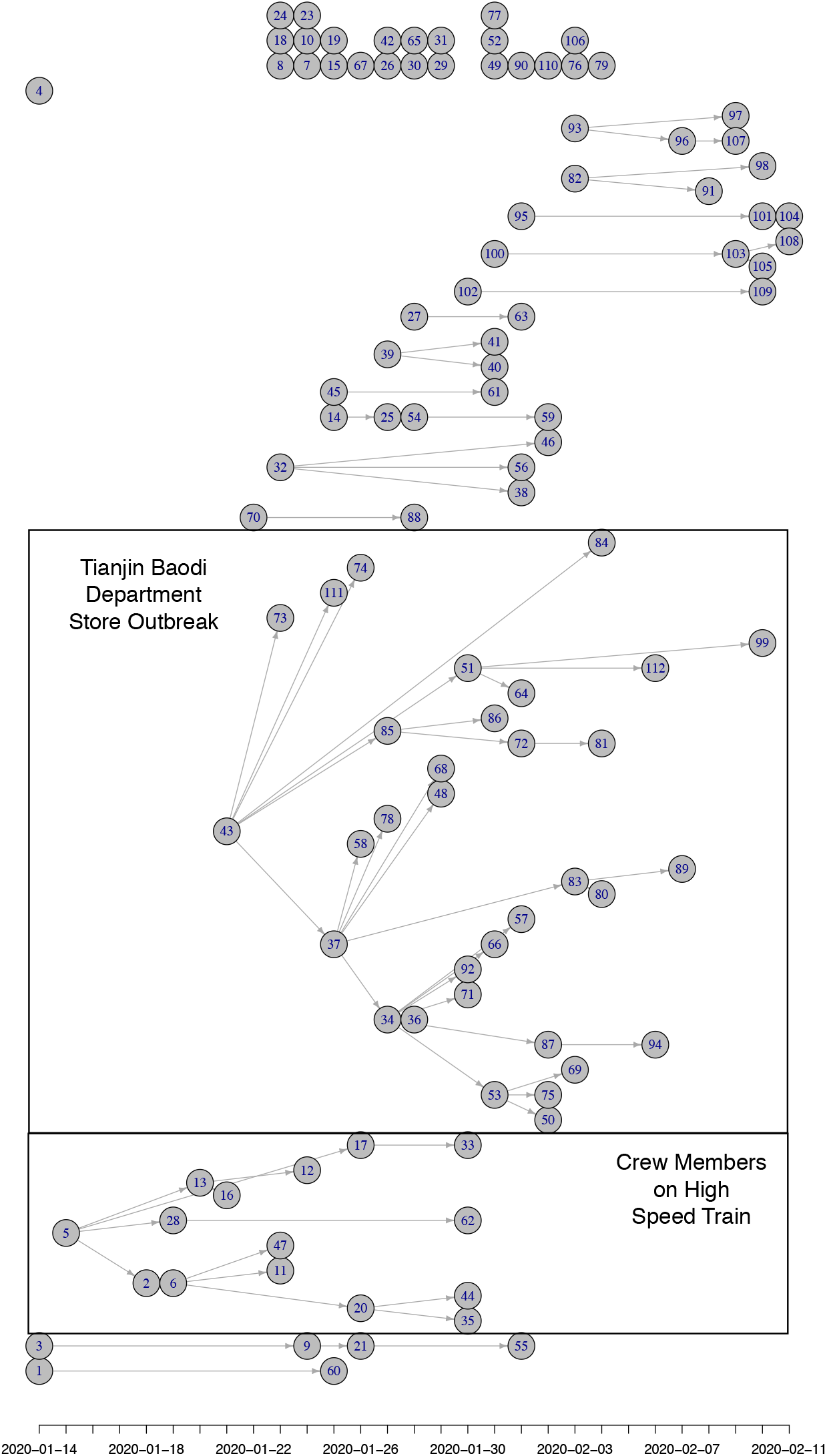
(Posterior mode) Transmission tree for Tianjin between 21 Jan 2020 and 12 Feb 2020. The x-axis represents the date of symptom onset. Most initial cases of clusters in Tianjin were imported from Wuhan/Hubei. There were two large clusters of cases: one among crew members of Tianjin railway and another one among sales representatives and customers in Baodi department store. Many small clusters were among relatives and friends and a proportion of imported cases did not infect anybody.

Figure 3 shows estimated reproduction numbers by day for the initial 425 confirmed cases in Wuhan. “The market” node as an infectious source was linked to as many as 13·6 cases, on average. For those 425 initial cases, the mean reproduction number was 2·5 (until 31 December 2019).

**Figure 3:**
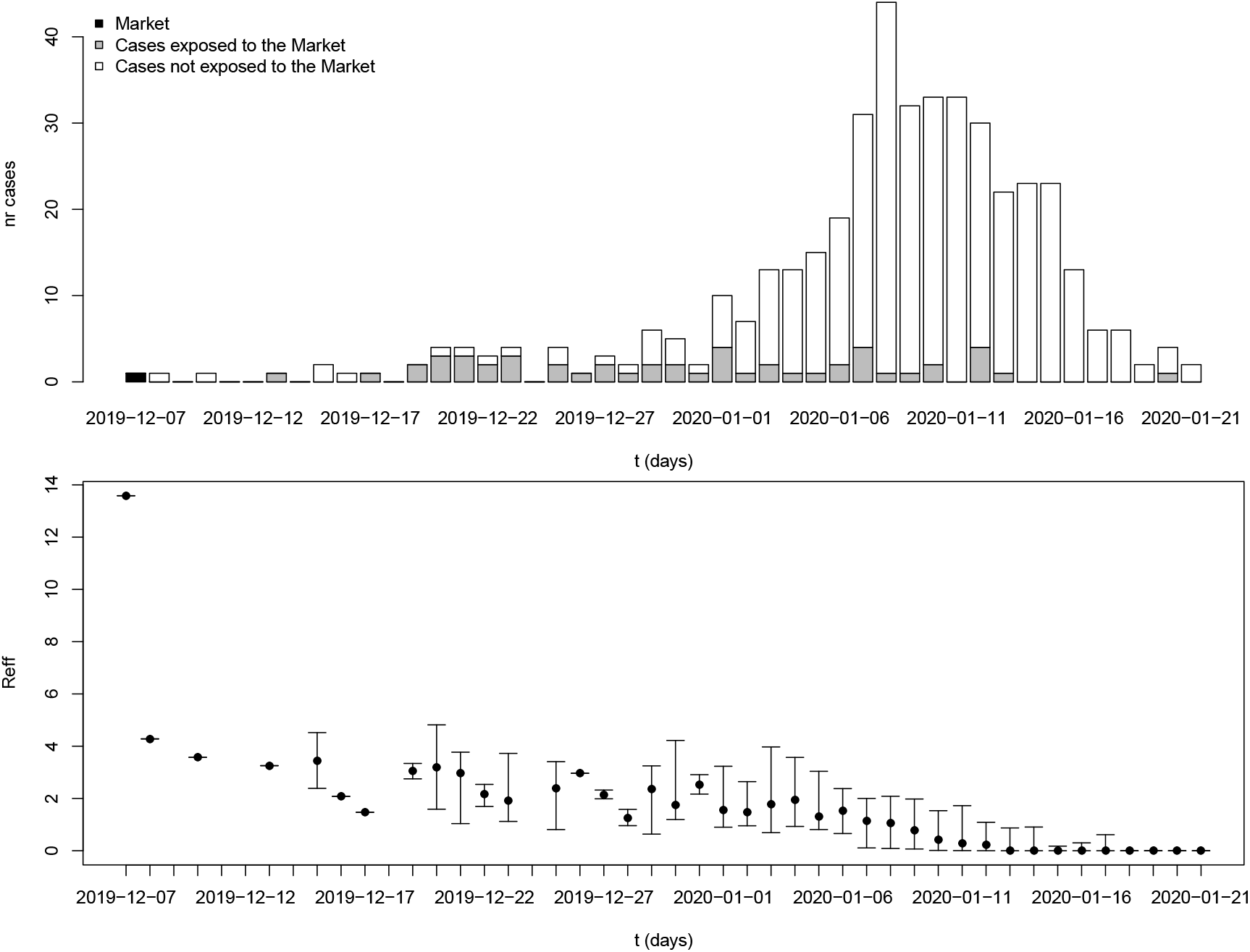
Reproduction number estimates for the period 8 December 2019 to 21 January 2020 in Wuhan, Hubei province.

The estimated numbers of imported cases from Wuhan varied among provinces in mainland China. Provinces bordering Hubei province include Henan, Hunan, Anhui, and Jiangxi (213, 175, 163, 146), and provinces with close economic ties with Hubei province include Zhejiang and Guangdong (271 and 228, respectively). These provinces all had high estimated numbers of cases imported from Wuhan by 3 February 2020 (Figure 4b).

**Figure 4:**
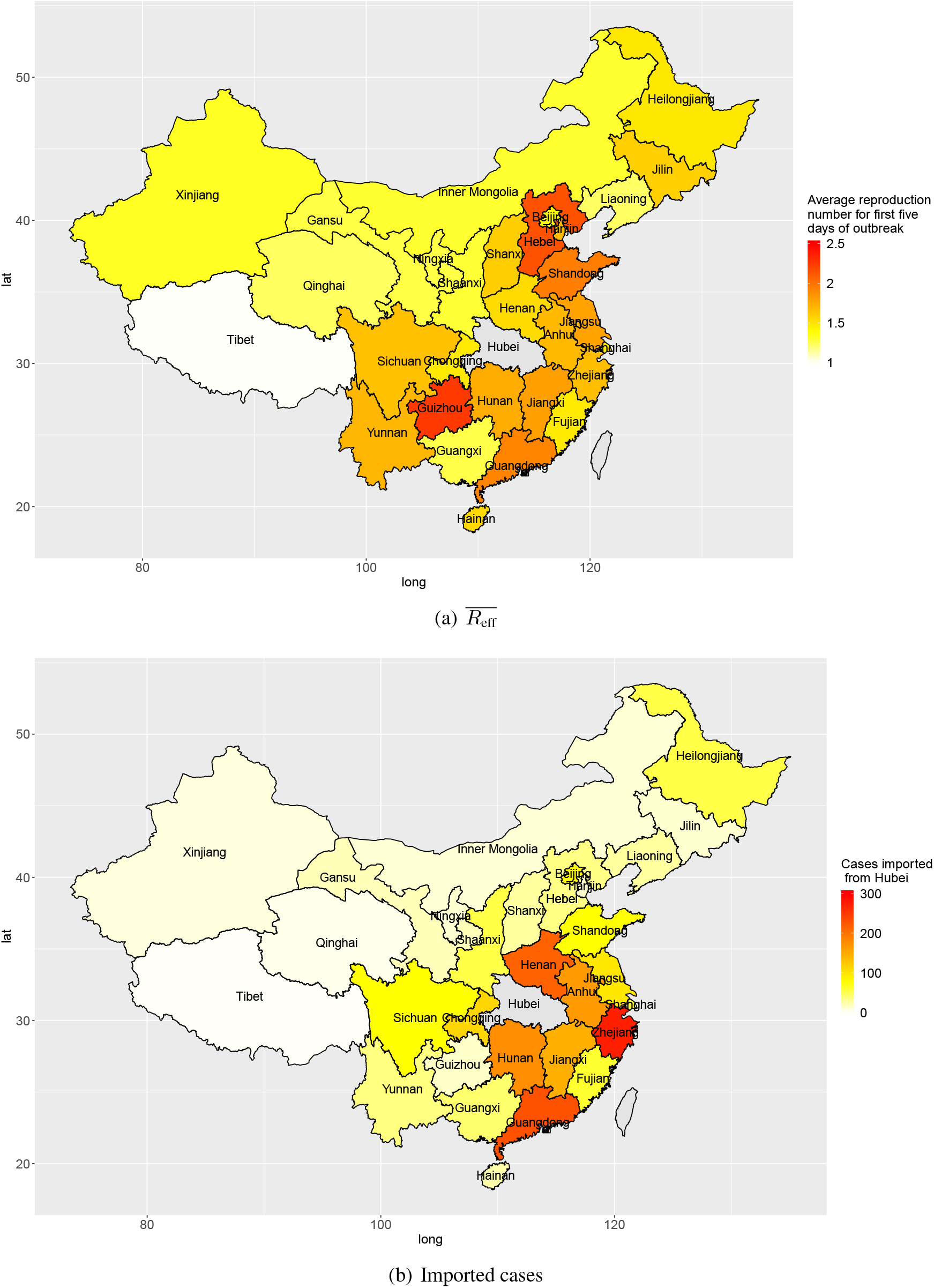
Transmission in provinces in China: average reproduction numbers (a) and estimated numbers of cases imported (linked to) Wuhan (b).

Outside Hubei there was less transmission than within Hubei province, with average reproduction numbers between 1 and 2 during the first five days after the first confirmed local case. The provinces Guizhou, Hebei, Shandong, and Guangdong had higher average reproduction numbers: 2·25, 2·14, 1·95, and 1·91, respectively (Figure 4a). Though the average reproduction numbers were not very high, there existed large variations in reproduction numbers of individual cases, which indicated some individual cases could have spread the disease to many people (Figure 2).

Within a province, there was also spatial variation in the amount of transmission. Figure 5 shows the disease transmission in 21 cities within Guangdong province. Shenzhen and Guangdong, the two largest and most affluent urban centers in Guangdong, had the highest estimated numbers of cases imported from Wuhan (127 and 124, respectively) by 3 February 2020. Guangzhou, Zhaoqing, Shaoguan, Shenzhen, and Zhuhai had reproduction numbers slightly larger than 1 (1·35, 1·31, 1·30, 1·10 and 1·05, respectively) for first five days after the first confirmation of a local case in the city.

**Figure 5:**
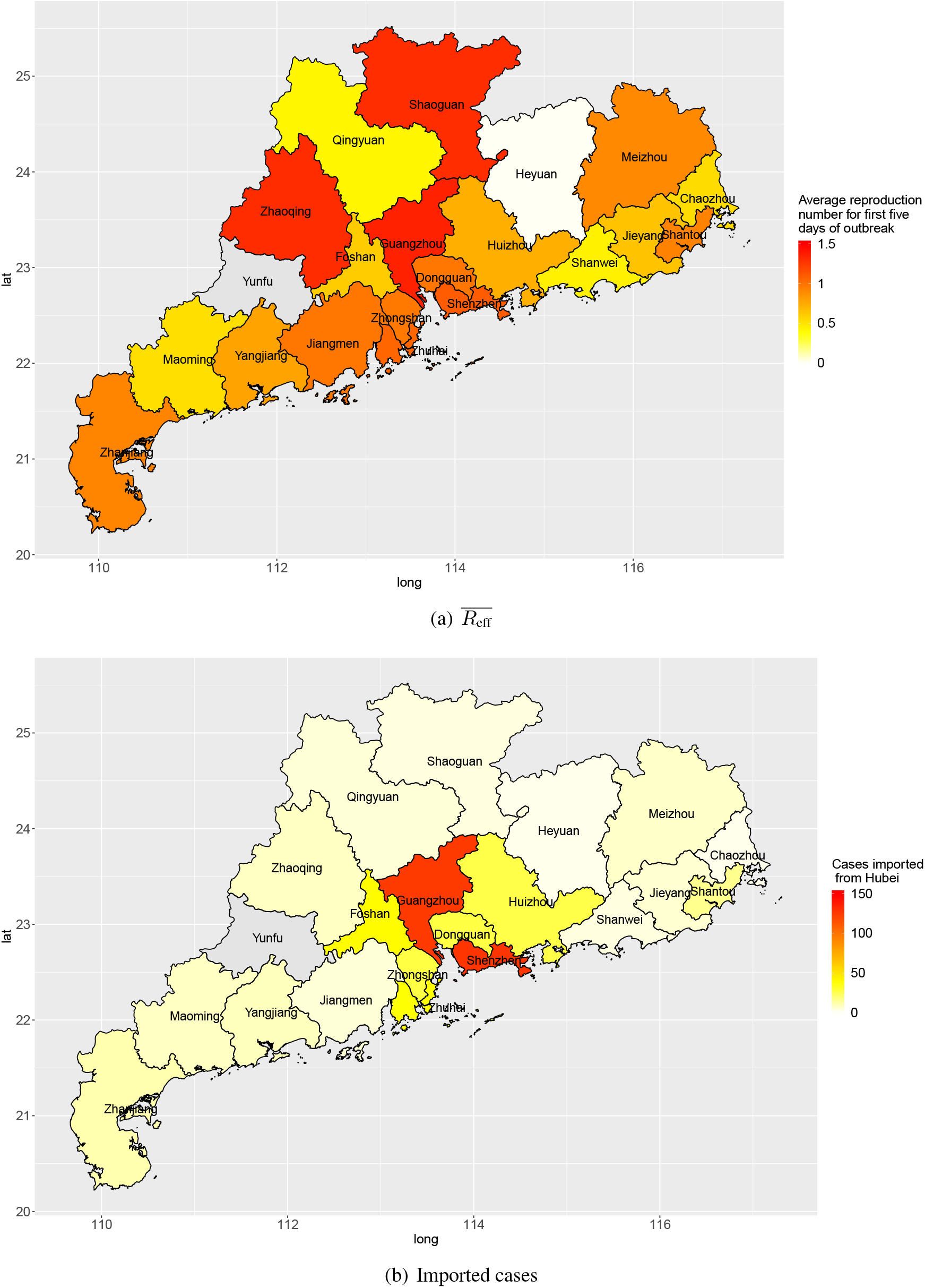
Transmission in cities in Guangdong province: average reproduction numbers (a) and estimated numbers of cases imported (linked to) Wuhan (b).

Figures A1 and A2 show boxplots of estimated reproduction numbers for provinces in China and cities in Guandong province, respectively. Figure A3 shows reproduction number estimates by gender and age for Beijing and Tianjin.

## Discussion

Spreading patterns of COVID-19 at different spatial scales, from local (railway department and department store) to regional (cities within Guangdong Province) to nationwide (Provinces within China), appear similar: new cases arise from contacts with a reservoir, spreading to remote locations through travel from the origin outbreak region. There likely is some secondary spread locally, from cases infected from the reservoir, but local reproduction numbers are low, often insufficient to support sustained transmission.

Knowledge of the serial interval, for symptomatic cases in the outbreak, is essential for analysis of the transmission probabilities [14]. Previous studies have estimated the distribution of serial intervals from a set of confirmed transmission pairs, where both the ancestor (who caused infection) and the descendant (who became infected) were known [19, 20]. As such information is mostly lacking [6] (and may be hard to obtain) in this COVID-19 outbreak, we have estimated the serial interval distribution from a curated network, using whatever contact information was available. Existing knowledge of contacts (and timing of those contacts) between cases was used to construct a matrix of prior probabilities, to restrict the transmission network to only those links that are possible.

When all transmission links are known, the serial interval distribution can be easily estimated. However, at early stages of an outbreak, information on transmission links is usually incomplete. When a sufficiently large proportion of the transmission links is known, it is possible to estimate both the serial interval distribution, and the transmission probability matrix [14]. Starting from a plausible serial interval distribution, the transmission probability matrix ***V*** is updated until convergence; then ***V*** is frozen and the serial interval parameters are updated, again until obtaining a new optimum; then the serial interval distribution is frozen and the transmission probability matrix is updated again, and so on, until no further improvement can be found. The estimated serial interval distribution, Gamma(3·16,1·52), has a mean of 4·8 days and good spread out (i.e. large scale parameter) to long serial interval. The variation of serial interval could caused by highly varied incubation period [21]. Such a short serial interval, compared to SARS (mean: 8·4 days) [22] and MERS (mean: 6·8 days) [23], gives COVID-19 ability to spread more rapidly. The rapid spread of COVID-19 in South Korea (from 31 cases on 18 February to 2,022 cases on 28 February) and Italy (from 20 cases on 21 February to 650 cases on 28 February), shows how missed infectious subjects may cause rapid transmission within a very short period, due to the combination of a short serial interval and an occasionally high reproduction number [24].

As the incubation period seems to be highly variable, it may be possible that appearance of symptoms in any case precedes symptom onset in its ancestor. When that happens, the serial interval for symptom onset is negative. To check whether negative serial intervals would adversely affect analysis we used an alternative distribution, where the serial intervals were shifted left-ward by a small amount. A shift of one or two days had no destructive effect on estimation of ***V***, and the resulting estimates of the effective reproduction numbers did not change substantially.

Early transmission of a newly emerging infectious disease in a population lacking immunity could reveal the basic reproduction number as long as there is no intervention. Currently, the source of COVID-19 is still unclear but many of the early cases were reported to have had contact with the Huanan Seafood Wholesale Market. A recent announcement released by the Chinese Academy of Sciences indicated that the source may not have come from this Seafood Market [25]. Any patient zero would have occurred before the initial 425 confirmed cases, but lacking any specific information about early spread of COVID-19, it can be assumed that a reservoir of infectious subjects could function as a super–spreading node causing new infections upon contact at any time within the outbreak. Assuming such a source for the initial cluster in Wuhan (“the Market”) led to an estimated basic reproduction number 13·6. It must be noted that this environmental “super–spreading node” is hypothetical, and represents the production of new cases by the joint presence of infectious subjects connected to the initial source.

There was strong heterogeneity in disease transmission in different provinces of mainland China. The present analysis produces two relevant characteristics: the numbers of cases imported from Wuhan, and the average reproduction number for the first five days after any confirmed imported case in each province. When there are many imported cases from Wuhan, these constitute a large base number of cases to start local spreading of the disease. Provinces geographically close to Hubei province are expected to experience mass migration from Wuhan, especially near the holiday season. Provinces with big tier–1 cities like Zhejiang and Guangdong both have very high estimated numbers of cases imported from Wuhan due to their economic connection with Hubei. When early transmission has a high reproduction number, local growth rate in a province will be high. Provinces geographically distant from Hubei, like Hebei, Guizhou, and Shandong, appear to have relatively high reproduction numbers during early local transmission. In provinces distant from Wuhan, the central hub of the outbreak, inhabitants and local governments may have been less cautious (with few public and personal prevention measures) considering high costs of interventions and perceived low risk.

Since Guangdong province has both large numbers of imported cases and high reproduction numbers for early transmission, we further examined the transmission inside of Guangdong by city. Shenzhen and Guangzhou, two of the biggest cities in China, had many imported cases. This is expected considering intense economic connectivity: transportation links, for goods and people including enormous numbers of migrant workers working in those two cities. The numbers of cases imported from Wuhan into other cities in Guangdong province was smaller. This pattern matches the flow of people in public transport (by train and airplane) converging onto the main transportation hubs (Shenzen and Guangzhou) and diverging from there towards other destinations in Guangdong province. For early transmission following import from Wuhan, the reproduction number in most cities in Guangdong had an average reproduction number around 1, during the first five days. With appropriate and sustained disease prevention and control measures (e.g. suspension public transportation, cancellation of mass gatherings, implementation of surveillance, and promotion of wearing face masks and personal hygiene), the outbreak is unlikely to spread out in those cities.

Among reported cases, gender differences seemed unimportant; most serious illnesses occurred among the elderly, in particular those with health problems prior to infection, while only a small proportion of cases were young of age [21]. In the present study, when reproduction number estimates were grouped by gender, no differences were found. Similarly, estimates of transmission from different age groups showed that small children and elderly people were equally likely to transmit infection as any other age group (Figure A3). We also examined the effect of the size of clusters imputed on the results and the difference was trivial between average cluster size three and five.

An important issue in analyzing transmission of COVID-19 is the amount of silent transmission. As mentioned earlier, some infectious subjects may transmit their infection before they become symptomatic. As long as their descendant cases also go on to develop symptoms their transmission link may still be established. However, when infected subjects who remain completely asymptomatic could be infectious to susceptible contacts, the appearance of such contacts, when symptomatic, could not be linked to their immediate ancestors. Given the fact that symptoms seem to be milder in those who are young, such unobservable transmission cannot be excluded [26]. Silent transmission has been seen in other infectious diseases [27] where, notably, asymptomatically infected subjects appeared to cause fewer transmission. As asymptomatic infections would lead to an antibody response, serology could be a valuable tool to assess the importance of asymptomatic transmission.

## Data Availability

Data used for this study are all from publicly available sources and
cited.

## Contributors

All authors contributed equally to this work.

## Data sharing

Data used for this study are all from publicly available sources and cited. The data files are available upon request to the authors.

## Acknowledgements

The authors received no specific funding for this work.

We are grateful to the alumni of cohort B5 ’08 at Shijiazhuang No.2 Middle School, including Di An, Jiayi Liu, Xun Li, Menghan Hou, Yunpeng Zhang, Fan Huang, and Hongxu Li, for gathering and sharing information form various sources about COVID-19.

## Supplementary material

**Fig. A1:**
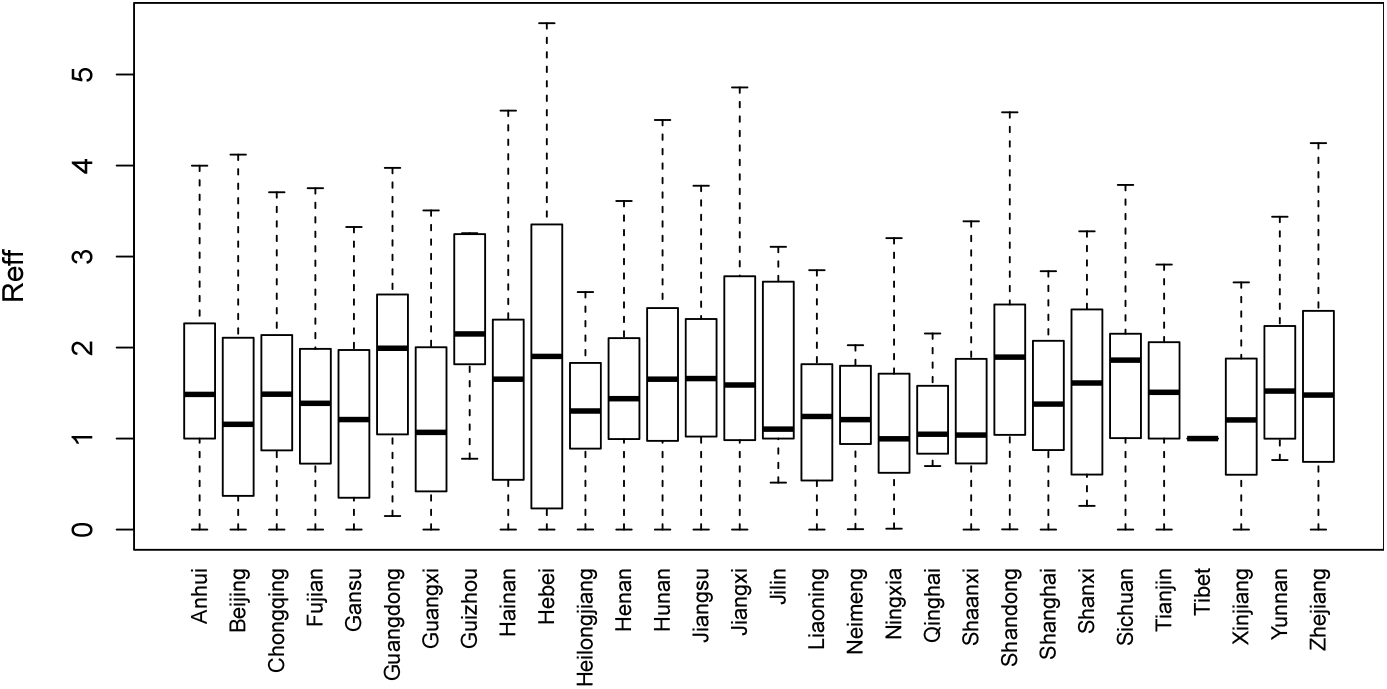
Reproduction number estimates for provinces in China.

**Fig. A2:**
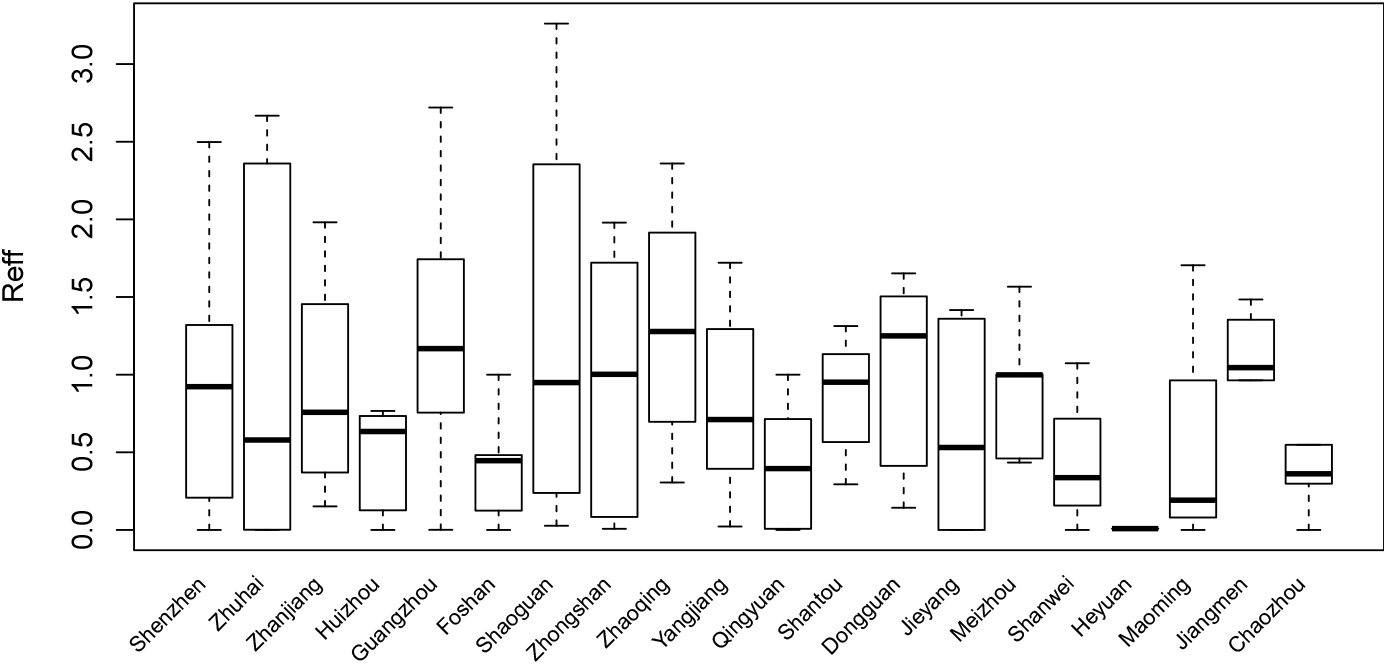
Reproduction number estimates for cities in Guangdong province.

**Fig. A3:**
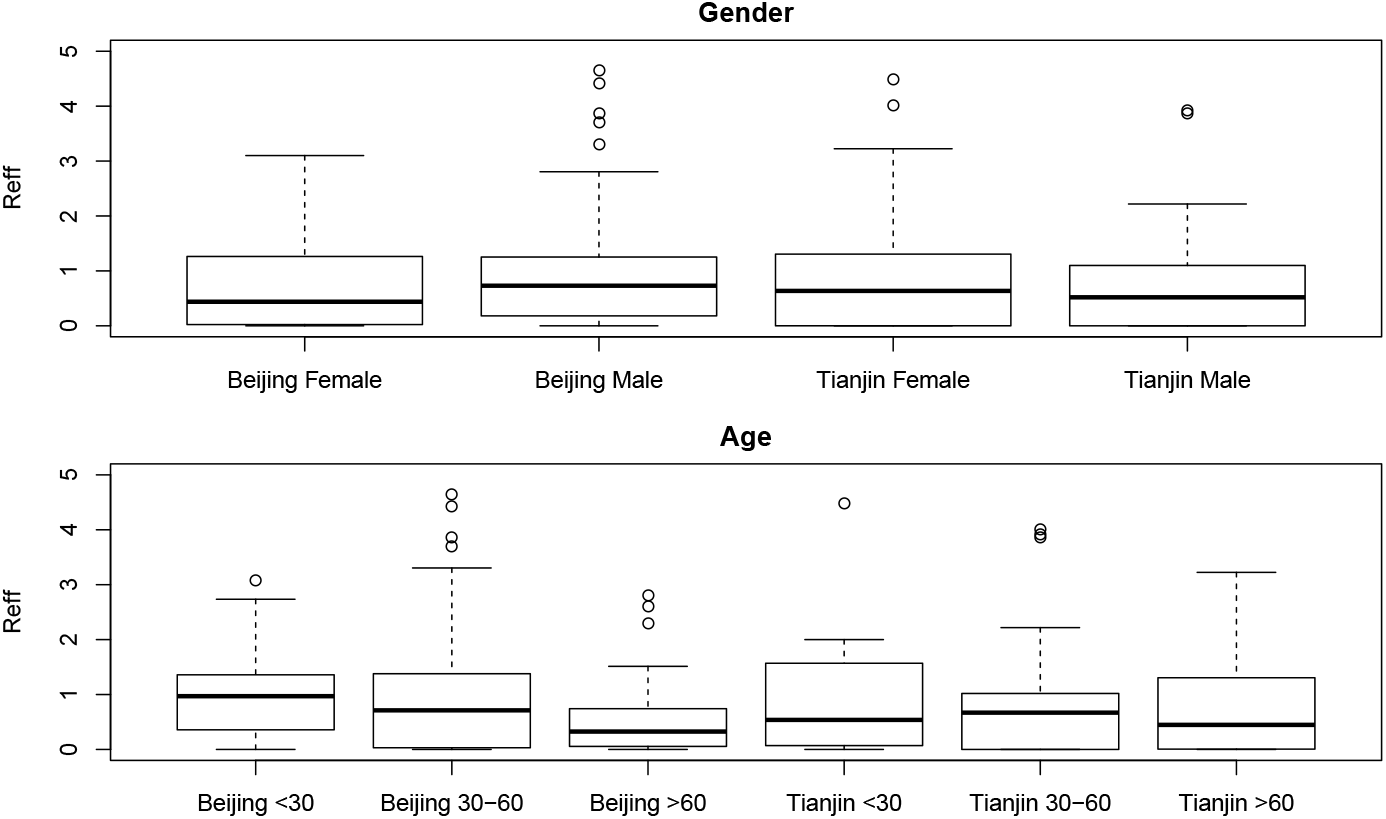
Estimated reproduction numbers by gender and by age category for Beijing (228 cases) and Tianjin (112 cases).

